# Detecting heterogeneous seizures in newborn infants using triple correlation

**DOI:** 10.1101/2023.06.09.23291216

**Authors:** Graham A Smith, Julia Henry, Wim van Drongelen

## Abstract

We detect seizures in newborn infants using a novel method derived from triple correlation, which integrates spatial and temporal structure in neonatal electroencephalograms (EEGs). Triple correlation natively encompasses analogues to a variety of lower-order approaches (auto-correlation, cross-correlation) in addition to introducing higher-order signals, so we hypothesized that our approach would both effectively detect and differentiate notoriously difficult-to-detect and heterogeneous neonatal seizures. Indeed, our method in its simplest form performs comparably well to a current standard of care, amplitude-integrated EEG (aEEG), and by some measures outperforms aEEG, suggesting at a minimum that a combination of triple correlation and aEEG could produce a more effective first-line bedside detector. Moreover, we find that the triple correlation seizure-signal varies between patients, with 1) differences in dominance of either within or between channel correlations and 2) differing levels of higher order structure. We hope that our approach will provide a fertile field for future work in distinguishing and detecting seizures.

## 1 Introduction

Newborn infants have the highest risk of seizures among all age groups, seizures that both often have severe consequences and cannot always be visually detected (Glass et al, 2016; Padiyar et al, 2020): only EEG monitoring can detect a majority of these seizures (Glass et al, 2013). Unfortunately, the gold standard, continuous video EEG (cEEG) monitoring, is highly resource intensive, requiring specialized clinicians to evaluate high-frequency data around the clock (Kubota et al, 2018). To improve efficiency, clinicians often use transformed and time-compressed EEG signals to improve the visual salience of seizures, which allows them to read through recordings more quickly (Glass et al, 2013). However, even with more efficient review, ongoing real-time monitoring is still unrealistic for the vast majority of neonatal care. While catching the seizure in review at the end of the day is helpful, the quicker the detection the better the outcomes (Gotman, 1985; Gotman et al, 1997). Ultimately, we would like a detector that could alert clinicians in real-time to the possibility of a seizure.

Such a detector might naturally be constructed using the same transformation that enables clinicians to more efficiently review recordings: an improvement in visual salience should correspond to an improvement in numerical distinguishability. In neonatal intensive care units (NICUs), the standard transformed signal is amplitude-integrated EEG (aEEG) (Glass et al, 2013). An aEEG signal includes two traces subsampling a rectified and low-pass-filtered EEG to include only two points in a large time window (e.g. 15 seconds): one at an upper percentile, one at a lower percentile (e.g. 95th and 9th) (Zhang and Ding, 2013; Rakshasbhuvankar et al, 2015; Chen et al, 2019). Since this subsamples a typical 256Hz EEG over 2000x, with aEEG clinicians can quickly review several hours of recordings to identify time windows with trends suspicious for seizure activity. Additionally, neonatologists lacking the specialized training required to read cEEG can parse aEEG bedside Glass et al (2013). One particular trend indicative of seizure is an increase in the lower trace, which is a trend in theory easily detectable by simple thresholding (Glass et al, 2013).

In this paper, we implement a simple detector with a signal derived from triple correlation. We have shown elsewhere that triple correlation completely characterizes any neural recording (Deshpande et al, 2023), so it should reflect any difference between seizure and non-seizure epochs. We show that a simple, theory-inspired summary of triple correlation leads to detection on par with the aEEG lower margin. Moreover, unlike the aEEG’s single axis of change, triple correlation reflects the heterogeneity of neonatal seizures.

## 2 Methods

### 2.1 Open-source EEG recordings

We used neonatal EEG recordings from an open-source dataset (Stevenson et al, 2019). These clinical recordings were sampled at 256 Hz and include evaluations by three expert reviewers for seizures. There were 79 patients in the dataset, with 343 seizures (defined by reviewer consensus) across 39 patients (seizing patient mean seizure count was 8.8 ± 11.9 std). The total duration of the recordings was 402,825s, of which 39,259s was marked as seizing (mean seizure duration 1,006s ± 1,392s std).

We ran our analyses both on the entire dataset, and on a subset with artifacts manually removed. Because of the time-consuming nature of expert artifact annotation, we only evaluated a subset of the recordings for artifacts. To choose that subset, we visually inspected all patients’ reviewer consensus plots (see Fig. 2) and selected eight patients as having both 1) good reviewer consensus, as indicated by a lack of epochs with only one or two of the three reviewers indicating a seizure, and 2) a good balance of seizure and non-seizure epochs. We then additionally took the first fifteen recordings by the dataset’s numbering, so that we evaluated a mix of “clean” and randomly-chosen recordings. A trained neurologist (JH) then annotated artifacts within these recordings. We discarded one patient (patient 21) from this subset because the vast majority of the recording was artifactual (92%).

**Fig. 1.**
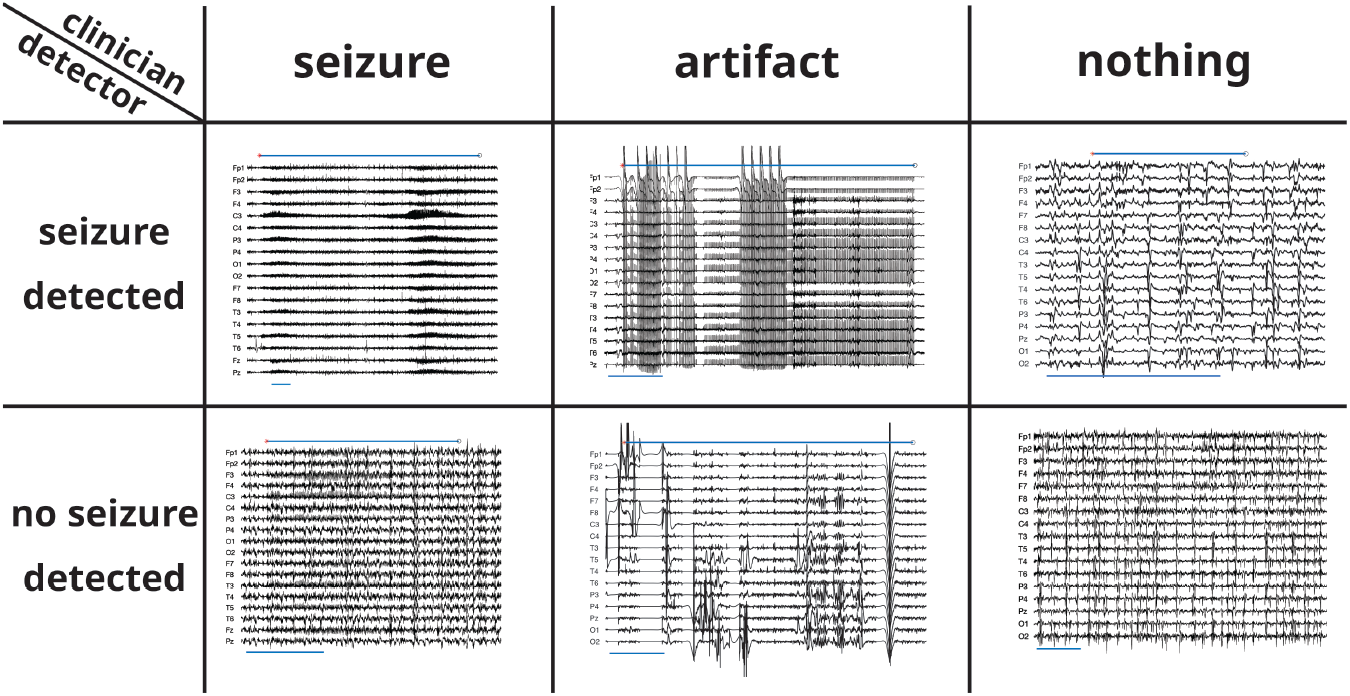
Example pre-processed EEG snippets in confusion matrix with artifacts. Snippets in the top row had salient TriCorr signals that were detected as seizures by most thresholds (see Fig. 3 for signal traces). Snippets in the bottom row did not have salient TriCorr signals. First-column snippets include a reviewer-consensus seizure (marked by upper bar). Middle-column snippets include a neurologist-annotated artifact (marked by upper bar). Third-column snippets include no expert-defined event, confirmed *post-hoc* by our neurologist (JH). The upper bar in the upper-right snippet indicates the erroneous detection. Green outlines indicate the detector matched the reviewer consensus, red indicates detector error. The blue bar below each recording indicates 60s.

**Fig. 2.**
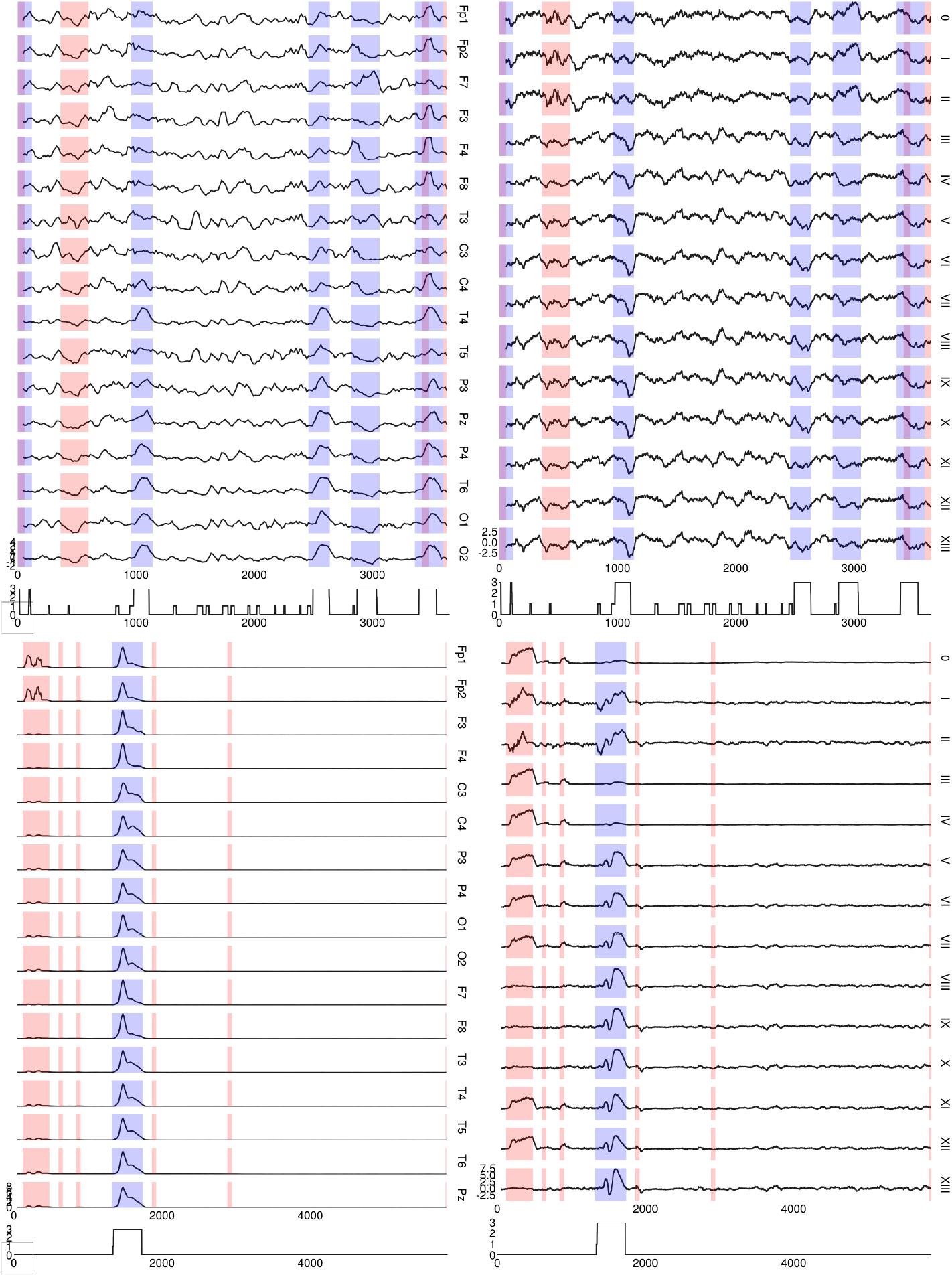
TriCorr and aEEG signals used in simple detector. **left** aEEG lower margins (9th percentile) for each EEG channel, for patients 7 (top) and 62 (bottom) **right** TriCorr contributions for each motif class within sequential 1s EEG snippets. The uncolored trace shows the number of reviewers indicating a seizure occured in each second, out of a possible three reviewers. Blue shading indicates reviewer consensus, plus one minute on either side. Red shading indicates minutes including seconds that were artifact-annotated by our expert reviewer

There were 22 patients in the artifact-annotated dataset, with 134 seizures (defined by reviewer consensus) across 16 patients (mean seizure count was 8.4 ± 11.9 std). Total duration of the artifact-annotated recordings was 121,533s, of which 2,898s were marked as seizing (mean seizure duration 929s ± 842s std).

We excluded the respiratory and ECG channels from all analyses because they are not EEG signals. We additionally excluded Fz and Cz from all analyses due to channel-specific whole-recording artifacts in many patients.

### 2.2 Code

The below analyses were conducted with [Julia](https://julialang.org/) v1.7.3 (Bezanson et al, 2017). All code is available on Github at grahamas/NeonateTriCorr. Filtering used the package JuliaDSP/DSP.jl. Statistical tests used JuliaStats/HypothesisTests.jl. ROC analysis used davidavdav/ROCAnalysis.jl.

### 2.3 EEG preprocessing

Prior to analysis, we preprocessed the EEG recordings. We first de-meaned per-channel; then bandstop filtered between 45 and 55 Hz with a sixth-order Butterworth (to account for European mains frequency of 50 Hz); then bandpass filtered between 0.1 Hz and 70 Hz with a second-order Butterworth; and finally divided by standard deviation, per-channel.

### 2.4 Amplitude-integrated EEG (aEEG)

To calculate the aEEG, we followed the example of Zhang and Ding (2013). Starting from the pre-processed EEG, we bandpass filtered between 2 and 20Hz using an FIR with a 50th order Hamming window, diverging from Zhang and Ding (2013) but in line with Werther et al (2017); Quigg and Leiner (2009). Next we computed the envelope by lowpassing below 0.32 Hz (corresponding to an RC circuit with a 500ms time constant, as in Zhang and Ding (2013)) with a 5th order Butterworth. Then, we divided the recording into sequential segments of length 15 seconds. Within each of these segments, we calculated the “margins”: the value at the 9th and 93rd percentiles, the lower and upper margins respectively. These margins constitute the aEEG trace. To construct our detector, we used only the lower margin, since a common feature of seizures is an increase in the lower margin (Glass et al, 2013). In contrast to prior work, we opted to include all channels rather than pick a single channel.

### 2.5 Triple correlation over time

We calculated the triple correlation as in Deshpande et al (2023), in one second snippets with lags in the ranges of -8:8 spatial and -25:25 temporal bins, using periodic spatial boundary conditions and data-padded temporal boundary conditions. With 17 EEG channels, a lag of 8 spatial bins encompasses the entire spatial domain. With a sampling frequency of 256 Hz, the temporal lags encompass approximately -98:98ms. We summarized the triple correlation by its fourteen motif classes, which partition the space of triplet motifs into qualitatively distinct classes embodying different computational patterns, e.g. synchrony, feedback, divergence, etc.

### 2.6 Distribution comparisons

We used an approximate two-sample Kolmogorov-Smirnov (KS) test to compare seizure and non-seizure snippets’ signals (TriCorr or aEEG), excluding non-seizure snippets within 60 seconds of seizure onset or offset. The KS test tests the null hypothesis that two samples could have been drawn from the same unknown distribution. Thus, significant p-values indicate that the seizure and non-seizure signals likely had differing statistical distributions.

### 2.7 Detector construction

For both aEEG and triple correlation (TriCorr) we obtained multi-channel signals: in the case of aEEG, these are the seventeen EEG channels, and in the case of TriCorr, these are the fourteen motif classes. Within these channels, we z-scored either within or across patients: we subtracted either the across-patient or within-patient channel mean and divided by the all-patient or within-patient standard deviation. Then we computed the rolling temporal mean in 60s windows. These channels are depicted in Fig. 2.

In the case of TriCorr, we then took the absolute value of these signals, since seizures could be reflected by either positive or negative deviations from the mean. In contrast, aEEG lower margin only reflects seizures through increases. We created a single timeseries by taking the maximum value across all channels.

To account for complications due to differing resolutions of TriCorr and aEEG (1s vs 15s snippets, respectively), we applied our detector per-minute: we divided the signals into 60s intervals and regarded any suprathreshold signal within an interval as a detection for the whole interval, and otherwise marked no detection for the interval. An interval was regarded as a seizure interval if all three reviewers marked a seizure within one minute of the start or end of the interval.

We depict examples of both aEEG and TriCorr detector signals in Figure 3.

**Fig. 3.**
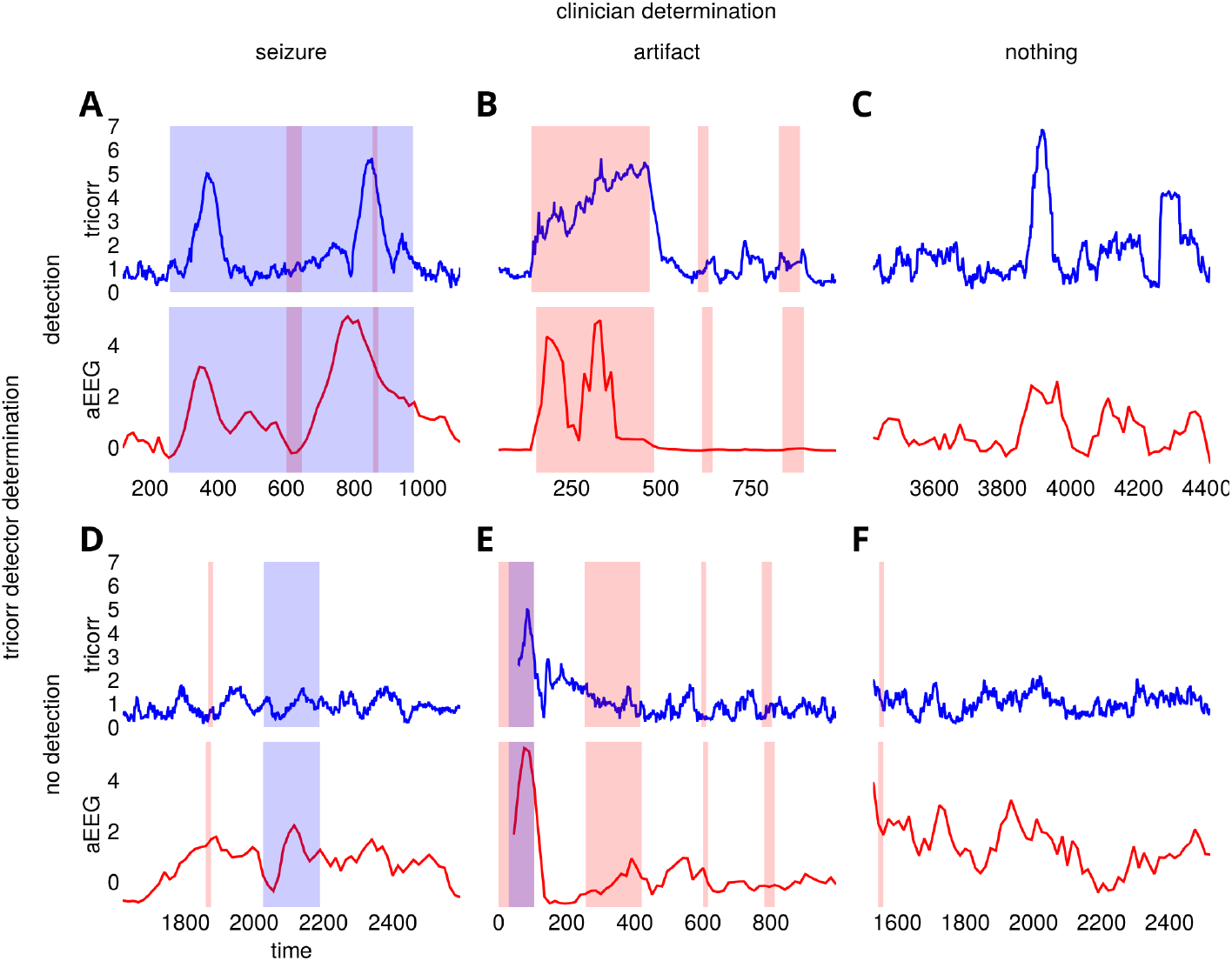
Example detector traces in confusion matrix with artifacts. Each trace is a 1000s long snippet including the example indicated by the row and column labels. These 1000s snippets encompass the same examples as in Fig. 1. We did not use a particular threshold to choose these examples. **A** true positives: a long seizure (blue highlight) with two high-amplitude detections by both TriCorr (blue line) and aEEG (red line). **B** artifact-induced false positive: three artifacts (red highlight), with the first producing salient signals in both TriCorr and aEEG. **C** a false positive in the TriCorr signal, though not so definitively in the aEEG. **D** false negative: a seizure not detected by either signal. **E** true negative (correct artifact rejection): the latter three artifacts are not marked as detections by either TriCorr or aEEG. However, the first artifact encompasses a seizure that is correctly detected by both TriCorr and aEEG, and so does not fall properly into any of these six categories. **F** a true negative with only a brief correctly rejected artifact.

### 2.8 Detector metrics

To evaluate the efficacy of our detectors, we constructed ROC curves using davidavdav/ROCAnalysis.jl. These ROC curves are depicted in Fig. 4 with clinically-relevant versions of the true positive rate (TPR) and false positive rate (FPR). Rather than a per-interval TPR, we reported a per-seizure TPR, which indicates the proportion of seizures correctly detected. For FPR, we reported the false positives per hour (FP/Hr), which is simply the FPR multiplied by the number of non-seizure hours. Note that AUCs are reported for the identically-shaped ROC but with a per-interval FPR axis rather than FP/Hr, so that the AUC is between 0 and 1.

**Fig. 4.**
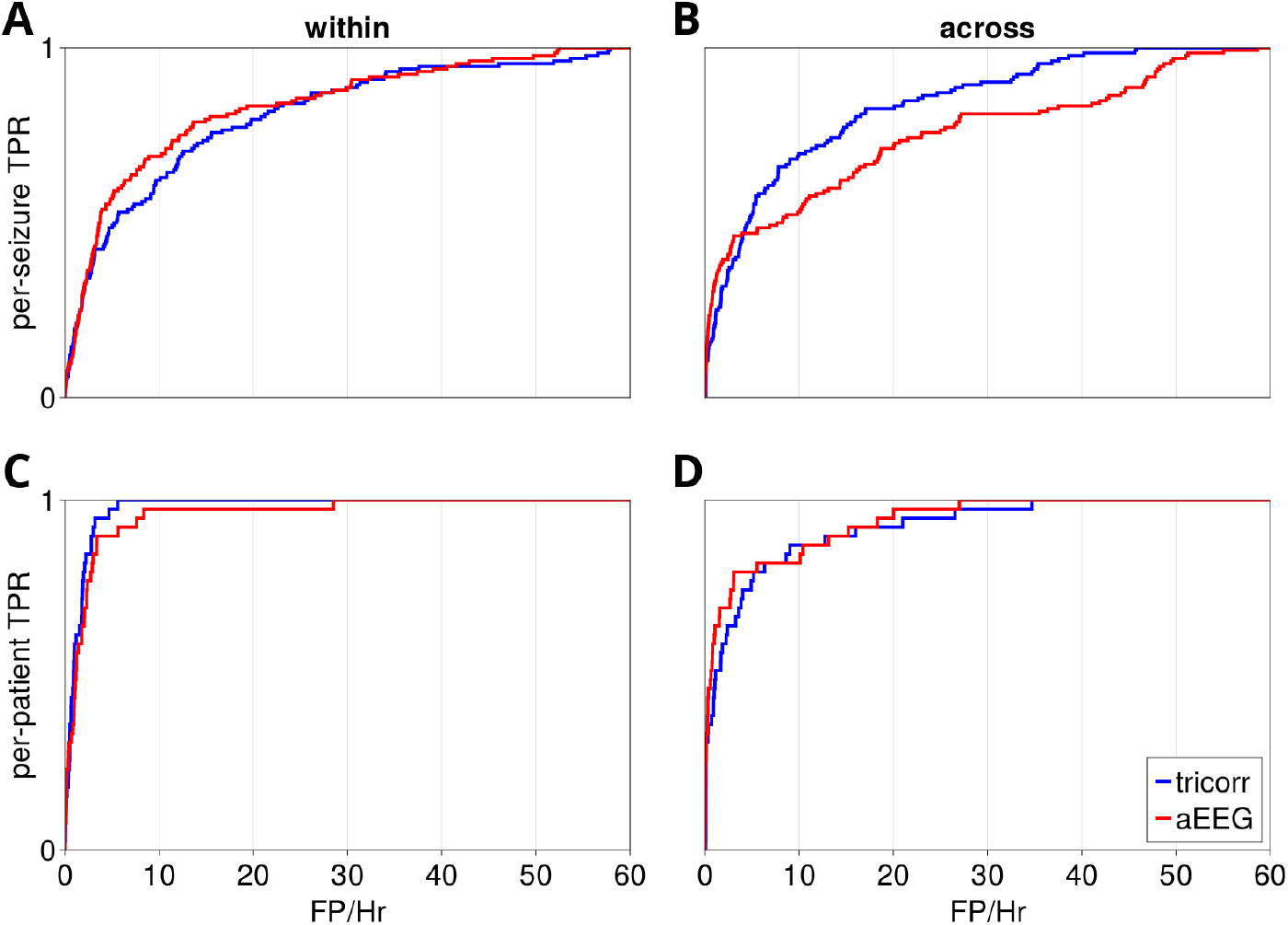
Detector ROC curves. These curves compare the success of detectors when defining success per-seizure, *top row*, or per-patient, *bottom row*, and when z-scoring the detector signals across all patients, *left*, or within each patient, *right*. In all cases the failure rate is defined as false positive minutes per hour.

We calculated the clinician FP/Hour rate by marking detections when *any* reviewer indicated a seizure and comparing this to the reviewer-consensus “ground truth.”

## 3 Results

### 3.1 Triple correlation detects individual seizures comparably well to clinical standard

A signal based on triple correlation leads to similar per-seizure detection performance compared to one based on aEEG, the current clinical standard. Neither is obviously better. The ROC curves are quite similar (Fig. 4), with the only substantial deviation being in the case where we z-score the signals across all patients for detecting individual seizures (Fig. 4B). In that case, the TriCorr detector has a higher AUC than the aEEG. This suggests TriCorr could be a better naive detector, i.e. that it would require less tailoring to the individual patient’s signal statistics.

Comparing traces in individual patients, in some patients the aEEG signal is cleaner, in others the triple correlation signal. Nor is it clear that the “ground truth” is any better: trained clinicians routinely disagree with each other. In our data, the trained clinicians had a false positive rate of 6 FP/Hr when treating single reviewers as the detection signal and reviewer consensus as the ground truth.

### 3.2 Triple correlation can detect seizing patients comparably well to expert reviewers

For per-patient detection, at perfect sensitivity the TriCorr detector can have the same specificity as the expert reviewers (Fig. 4C; Table 1), in the most ideal case where we have an estimate for the distributions of motif-class contributions for each patient. Given that this level of uncertainty is inherent in our “ground truth,” this performance is the best performance we could expect. In comparison, even in this ideal case aEEG does not achieve perfect sensitivity without more than five times the reviewers’ FP/hour. The aEEG’s sensitivity at the same 5.7 FP/Hr specificity is 92% (see Table 1).

**Table 1.** Statistics for the ROC curves shown in Figure 4. Target indicates whether the positives were per-patient or per-seizure; standardization indicates whether the signals were z-scored across or within patients. Signal indicates aEEG vs TriCorr. AUC is area under the ROC curve. TPR indicates the true positive rate (either per-patient or per-seizure) at FP/Hr = 5.7, which was chosen because that is the false positive rate of single-reviewer annotation. FP/Hr indicates the rate of false positives for a detector with 80% sensitivity.

| Target | Standardization | Signal | AUC | TPR (5.7 FP/Hr) | FP/Hr (80% TPR) |
| --- | --- | --- | --- | --- | --- |
| patient | across | aEEG | 0.94 | 0.82 | 5.5 |
| patient | across | tricorr | 0.92 | 0.79 | 6.3 |
| patient | within | aEEG | 0.96 | 0.92 | 2.9 |
| patient | within | tricorr | 0.98 | 1.0 | 2.1 |
| seizure | across | aEEG | 0.76 | 0.48 | 27.0 |
| seizure | across | tricorr | 0.84 | 0.58 | 16.0 |
| seizure | within | aEEG | 0.83 | 0.6 | 15.0 |
| seizure | within | tricorr | 0.81 | 0.52 | 21.0 |

### 3.3 Triple correlation distinguishes heterogeneity

Triple correlation reveals structural variations between patients’ seizure responses, where aEEG only offers a monolithic signal. Our analysis reflects that neonatal seizures can involve either an increase or a decrease in triple correlation, and usually this change is consistent within a patient, i.e. each patient has a particular type of seizure. Our algorithm then reduces this complexity to ask if signals indicating any of these types of seizures are present. However, heterogeneity in seizure signal is visually apparent before this reduction Fig. 2.

Of the 39 patients with seizures, 13 had significant distribution differences with increases in triple correlation ictal vs interictally, 11 had decreases, 3 had heterogeneous effects among motif classes, and 11 had no significant difference in motif class contributions between ictal and interictal snippets, as determined by a Kolmogorov-Smirnov test.

## 4 Discussion

Triple correlation provides a rich field of potential signals. Here we used only the simplest and even so matched and perhaps exceeded aEEG, a signal transformation in wide clinical use. Even better, our approach distinguishes among the heterogeneous neonatal seizures, even here where we have reduced TriCorr to its grossest components (motif classes). In its fullness, triple correlation provides a plethora of building blocks with which to construct signals with the potential to differentiate between seizures. Perhaps we will one day be able to relate these differences to known differences in etiology and treatment.

The expressiveness of TriCorr signals could also help more immediately, at the bedside where aEEG is currently used. Commonly aEEG is used as a front-line bedside tool by neonatologists rather than neurologists because it offers a straightforward signal that doesn’t require years of training the way that cEEG does. However, that simplicity comes at a tradeoff when some nuanced differences in signals have different diagnostic outcomes. For example, one critical predictor of neonatal outcomes is the EEG background, which can be read from the aEEG margins over an extended time. However, abnormal background voltage involves a decreased lower margin, which stands in contrast to seizure detection, where an increased margin indicates a seizure. This can lead long seizures to be mistakenly interpreted as normal background EEG, with concommitant incorrect treatment, and hence poor outcomes (Glass et al, 2013). With the expressiveness of TriCorr, we hypothesize seizure and normal background would be separable.

The further reaching advantage of TriCorr is that it specifically expresses features that are more covert than those expressed by aEEG. aEEG makes overt the general trends of the maxima and minima of the EEG, less some noise. In contrast, TriCorr overtly represents the third-order structure. While an expert could in theory read such structure from the EEG, it would be with extensive training and experience, not unlike that of the trained epileptologist. TriCorr creates the potential to explicitly represent that structure to a more generally trained audience, such as a nurse or neonatologist, for whom noticing a seizure is but one of many tasks.

This brings us to the final caveat of this study: we do not claim this will replace the need for neurologists trained in the art of reading cEEG. We can only argue that this prepares the way for more reliable and timely alarm to bring cases to the attention of the neurologist for definitive diagnosis. We do, additionally, hope that TriCorr will augment the neurologist’s reading, but this study does not begin to provide evidence for such a claim, merely the hope. Indeed, it is hard to see how such evidence could be acquired definitively, given that the ground truth must always be the expert reviewers themselves.

## Data Availability

The neurologist artifact annotation of the source data is available within our Github repository. All figures can be reproduced using our Github repository in combination with the publicly available source data.

https://github.com/grahamas/NeonateTriCorr

## Acknowledgments

G.S. is supported by the Pritzker Endowment for the Neurosciences.

## Statements and Declarations

The authors have no relevant financial nor non-financial interests to disclose.

